# Air pollution exposure when cooking with electricity compared to gas

**DOI:** 10.1101/2023.04.10.23288249

**Authors:** Carlos F. Gould, Lissete Dávila, M. Lorena Bejarano, Marshall Burke, Darby W. Jack, Samuel B. Schlesinger, José R. Mora, Alfredo Valarezo

**Author notes:** These authors contributed equally (Carlos F. Gould, Lissete Dávila). Communicating author: Carlos F. Gould; Stanford University, Department of Earth System Science; 473 Via Ortega, Stanford, California 94305.

## Abstract

We report small-sample evidence from a randomized experiment among a set of urban Ecuadorian households who owned both electric induction and gas stoves. We randomly assigned households to cook only with one stove during a prescribed two-day monitoring period, and then cook only with the other stove in a subsequent two-day period. The order of stove use was randomized, and air pollution was measured during each period. We found that mean 48-hour personal NO_2_ exposure was 9.9 ppb higher (95% CI, 4.5-15.3) — a 50% increase over the 48-hour induction mean — when households were randomized to gas as compared to induction. Mean kitchen area NO_2_ concentrations were 1 ppb higher (95% CI, 0.4-2.1) (a 6% increase) and mean personal PM_2.5_ exposure was 11 *μgm*^−3^ higher (95% CI, -0.1-22.8) (a 44% increase) during study periods when randomized to gas. We use time-resolved cooking and pollution data to illustrate that these differences are driven by LPG cooking, which was associated with a 5.0 ppb increase in 5-minute average NO_2_ kitchen area concentrations (95% CI, 3.4-6.7) and a 20.8 *μgm*^−3^ increase in 5-minute average personal PM_2.5_ exposure (95% CI 8.9-32.6). In contrast, cooking with induction was not associated with changes to short-term NO_2_ kitchen area concentrations, though it was associated with short-term increased personal PM_2.5_ exposure (10.8, 95% CI, 5.7-15.9).

## Introduction

The contribution of gas cooking to indoor air pollution and health risk is important for informing policy and personal choices in both industrialized and industrializing countries, but is poorly quantified. For roughly one-third of the world who cook with biomass daily, switching to gas could reduce air pollution exposure.^1^ However, if cooking with gas also poses health risks, electric stoves that produce no in-use emissions may be a promising ‘leapfrog’ technology that could yield both health and climate benefits, as promoted in recent household electrification initiatives.^2, 3^

Nitrogen dioxide (NO_2_) exposures are of particular concern when considering gas cooking as an environmental health hazard given previous cross-sectional evidence documenting higher NO_2_ concentrations in households using gas stoves as compared to electric ones,^4, 5^ and evidence that NO_2_ is causally related to poor respiratory outcomes.^4^

Among a set of urban Ecuadorian households who owned both electric induction and gas stoves, we randomly assigned households to cook only with one stove during a prescribed two-day monitoring period, and then cook only with the other stove in a subsequent two-day period. The order of stove use was randomized, and air pollution measured during each period. This study design afforded two key analytic advantages over previous cross-sectional studies. First, participants served as their own controls, which eliminates time-invariant confounding factors like kitchen characteristics and cook-level behaviors. Since randomized interventions replacing gas with electric cooking remain rare,^6^ existing evidence comparing households that decide to use different stoves may be biased by unobservable factors that drive both pollution differences and stove choices between households. Second, participants were already familiar with both cooking technologies, having used gas and induction for cooking for an average of 30 and 4 years; this alleviates common concerns in randomized interventions with short follow-up periods where households must adapt to a new technology and thus might not be generalizable.

## Materials and Methods

### Measurements

Data collection occurred between March 2021 and December 2021 in Quito, Ecuador. Primary cooks (*N* = 38) were randomly assigned to cook with only gas or induction in the first 48-hour period and then were instructed to cook with the other stove in a subsequent 48-hour period. Our primary outcome was time-integrated personal NO_2_ exposure, measured using passive badges (OGAWA PS-100) affixed near the breathing zone of a monitoring vest that participants were instructed to wear at all times except when bathing and sleeping. Monitoring vests also had a time-resolved, light-scattering personal fine particulate matter (PM_2.5_) exposure monitor (PATS+). Twelve participants were randomly selected to form an “intensive” monitoring group that included gravimetric PM_2.5_ (Ultrasonic Personal Air Sampler) and time-resolved kitchen area NO_2_ concentrations (AeroQual Series 500); half of this subset had duplicate NO_2_ passive badges, which were averaged in analyses. Time-resolved stove use was determined for the assigned stove using temperature loggers and current-voltage meters for the LPG and induction stoves, respectively. Cooking was identified based on highly positive slopes over short periods of time and when > 40 degrees C or > 2.5 V; cooking and non-cooking events had minimum durations of 5 and 30 minutes, respectively.

### Statistical Analysis

We estimated the effect of stove randomization on pollution in panel fixed effects (FE) regressions via ordinary least squares (OLS). The exposure was randomized stove assignment (reference: induction). The outcome was, separately, 48-hour average personal NO_2_ exposure, kitchen area NO_2_ concentrations, and personal PM_2.5_ exposure. We included FE (i.e., separate intercepts) for participant, month of year, and day of week. This intention-to-treat (ITT) analysis can be considered a lower bound of the estimate of the effect of gas cooking on pollution, since any deviation from stove assignment would attenuate the true effect.

We conducted some robustness checks. First to account for potential background pollution variations, we controlled for average ambient air pollution (NO_2_ or PM_2.5_) during each monitoring period from the nearest central site monitor (typically in the same neighborhood). Second, we controlled for potential variation in pollution monitor wearing by including the fraction of observations between 6am–10pm where PATS+ movement is detected. Third, we estimated the effect of the treatment on the treated by dividing our ITT estimate by the average fraction of total minutes cooked on the assigned stove when both stoves were monitored at the same time.

We estimated the effect of cooking events on short-term changes to kitchen area NO_2_ concentrations and personal PM_2.5_ exposure (both rolling 5-minute means) using panel FE regressions estimated via OLS where the exposure was whether LPG or induction stove use was detected, modeled separately and jointly. Here, we included FEs for participant, month of year, day of week, hour-of-day, and, for kitchen NO_2_, monitor FEs.

Standard errors were clustered at the participant level and *α* = 0.05 was used to determine statistical significance everywhere.

### Ethics

Research was approved by the Institutional Review Boards at the Columbia University Medical Center and the Bioethics Committee at the Universidad San Francisco de Quito (USFQ). USFQ approved COVID-19 safety protocols for in-person activities. Participants provided informed written consent online prior to visits or provided informed written consent on the day of visits. After the study, participants were given six beverage glasses or a pitcher and were provided with a summary of their cooking and exposures.

## Results

Household characteristics are summarized in Table S1. Air pollution measurements, detected cooking events, and detected monitor wearing are summarized in Table S2. Adherence to randomization was high: participants almost exclusivey used the assigned stove during the designated period and minutes cooked were comparable across randomization (Table S2, Figure S2).

Mean 48-hour personal NO_2_ exposure was 9.9 ppb higher (95% CI, 4.5–15.3) when households were randomized to gas — a 50% increase over the induction mean (Figure 1, Table S3). Half of induction period exposure estimates fell below the World Health Organization 24-hour NO_2_ guideline (13.29 ppb)^7^ as compared to one in ten in the gas period. Mean kitchen area NO_2_ concentrations were 1.1 ppb higher (95% CI, 0.4–2.1) (6% higher) and mean personal PM_2.5_ exposure was 11.4 *μgm*^−3^ higher (95% CI, -0.1–22.8) (44% higher) when randomized to gas (Table S4, S5).

**Figure 1:**
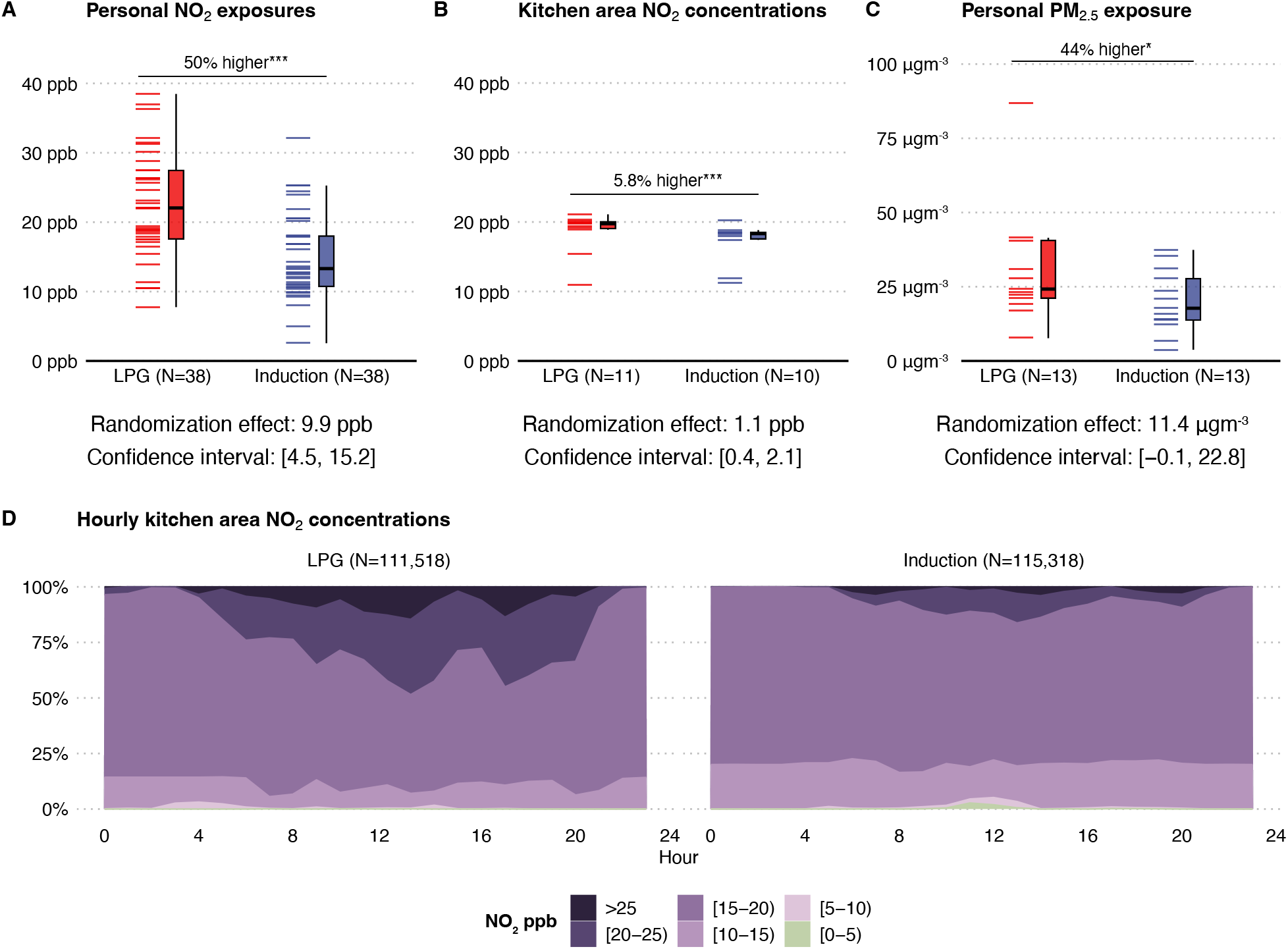
Personal and kitchen area air pollution concentrations when participants were instructed to use their LPG or induction stove exclusively and effect of randomization on pollution. In panels A–C, percent differences in means are estimated as based on the induction group average and estimated effect of randomization from the intention-to-treat analysis, which is noted below each panel along with the 95% confidence interval. Stars indicate statistical significance were P<0.10 = *, P<0.05 = **, P<0.01 = ***. (**A**) Shows integrated individual 48-hour personal NO_2_ exposures from passive badges and summarizes them in box-and-whisker plots according to which stove participants used. Induction *N* = 38; LPG *N* = 38. (**B**) Summarizes 48-hour kitchen area NO_2_ concentrations averaged from time-resolved data and summarizes them in box-and-whisker plots according to which stove participants used. Induction *N* = 10; LPG *N* = 11. (**C**) Shows integrated individual 48-hour personal PM_2.5_ exposures from integrated gravimetric filter data and summarizes them in box-and-whisker plots according to which stove participants used. One observation in the LPG group is not shown and was removed from analysis based on implausibility (mean = 513 *μgm*^−3^). Induction *N* = 13; LPG *N* = 13. (**D**) Shows the distribution of short-term (5-minute) kitchen area NO_2_ concentrations grouped by hour of the day and stove used. Minute-resolved observations: Induction *N* = 115, 318; LPG *N* = 111, 518

Detected LPG cooking was associated with a 5.0 ppb increase (95% CI, 3.4–6.7) in 5-minute average NO_2_ kitchen area concentrations and a 20.8 *μgm*^−3^ increase (95% CI, 8.9–32.6) in 5- minute average personal PM_2.5_ exposure (Table S6, S7). Cooking with induction was not associated with changes to short-term NO_2_ kitchen area concentrations, but was associated with increased personal PM_2.5_ exposure of 10.8 *μgm*^−3^ (95% CI, 5.7–15.9) (Table S6, S7).

Results were robust to inclusion of device wearing and ambient air pollution as controls and alternative specifications (Tables S3, S4, S5). The effect of treatment on the treated for 48-hr personal NO_2_ exposure was 10.3 ppb higher when households were randomized to gas, quite close to our ITT estimate.

## Discussion

Limitations of this study include its small sample size, which was budget constrained. Our confidence intervals were wide and future studies could benefit from recruiting more participants and repeating the procedure multiple times during a year. While our study has strong internal validity, our findings may not generalize to other settings (e.g., high-income countries).

We estimate an increase of roughly 10 ppb in 48-hour average NO_2_ exposure when participants were randomized to use the gas stove as compared to induction — similar to existing evidence in levels and percent difference, though not directly comparable since most studies are cross- sectional and do not measure personal exposures.^4–6^ Based on existing global evidence, this difference implies a 6% increase in risk of current and lifetime asthma^8^ or a 23% increase in risk of developing childhood asthma between birth and 18 years if differences are considered to be long term^9^ or, in the short-term, a 0.85% increase in risk of next-day mortality.^10^

We provide small-sample, high-quality evidence that gas cooking is associated with higher NO_2_ exposures relative to cooking with electricity. While similar evidence has been accumulating since the 1970s, better quantifying the health co-benefits of replacing gas with induction cooking could provide further support for residential electrification efforts, as well as for individual household health-protective choices. Randomized or quasi-experimental studies at the household level with longitudinal follow-up on cooking and health are recommended to provide policy- and health-relevant causal estimates of the benefits of replacing gas with induction stoves.

## Data Availability

Code will be available upon publication. De-identified in the present study are available upon reasonable request to the authors.

## Acknowledgments

The authors acknowledge funding support from the US National Institutes of Health Common Fund through the Clean Cooking Implementation Science Network. The authors are grateful to Dr. Sam Heft-Neal for helpful figure edits and thoughtful comments from Misbath Daouda, Dr. Minghao Qiu, and members of the Stanford ECHO Lab.

## Supplemental Information

### Details on NO2 passive samplers

We used standard UV–visible spectrophotometry (Ogawa 2006) to analyze the passive samplers. We measured temperature and relative humidity during each sampling with a co-located sensor to assist in calculating final NO2 concentrations. We calculated the limit of detection (LOD) as the mean plus three times the standard deviation of concentrations among blanks. We estimated an LOD of 1.9 ppb, similar to the other studies (Kephart et al. 2021) and the manufacturer recommended lower range of accuracy (2 ppb) (Ogawa 2006). The obtained determination coefficient for the calibration curve was 0.9997.

### Summary of air pollution measurements

Mean 48-hour personal NO_2_ exposure was 19.8 ppb (*n* = 77, standard deviation [SD], 12.2), mean 48-hour kitchen area NO_2_ concentration was 14.1 ppb (*n* = 15, SD, 6.1) and mean 48-hour personal PM_2.5_ exposure was 25.0 *μgm*^−3^ (*n* = 28, SD, 15.6) (Table S2). In paired observations, mean 48-hour personal NO_2_ exposures were 4.0 ppb higher (95% confidence interval [CI], 0.3–7.7) than mean 48-hour kitchen area concentrations.

Nighttime pollution measurements were similar across stove assignments (Figure S3).

We observed deviations from assigned stove use during 4.4% of total cooking time when LPG and induction stove use were measured (272 out of 6243 minutes across 15 monitoring sessions). In most cases, this was detected LPG use during the induction stove use monitoring sessions. In some cases, this was both stoves being used at the same time.

### Estimating increased risk from observed exposure differences

#### Child asthma

Khreis et al. conducted a meta-analysis of 21 studies and generate a summarized effect estimate of an odds ratio of 1.05 (95% CI 1.02–1.07) per exposure to 4 *μgm*^−3^ NO_2_ for the development of asthma at some point between birth and 18 years of age, i.e., an increase of 5% in risk. First, we translate *μgm*^−3^ to ppb, assuming 1 atmosphere of pressure and 25 degrees Celsius temperature, using a conversion factor of 1.88 *μgm*^−3^ = 1 ppb. Thus, 4 *μgm*^−3^ = 2.13 ppb. We then divide our observed effect estimate of 9.881 ppb by 2.13 ppb and multiply this difference by 5% (from OR = 1.05). Thus, we get an estimated increase in risk of developing asthma before 18 years of age of 23%. A limitation of using this effect estimate is that the included studies are primarily from outdoor air pollution. It is not immediately clear based on existing evidence how an increase of one unit outdoor air pollution will affect personal air pollution exposures. It is plausible that an increase of one unit outdoor air pollution will affect personal exposures somewhat less, i.e., less than one unit increase in personal exposure. In this case, we would expect an increased risk estimate of 23% to be an underestimate.

As a point of comparison, we can also draw on Lin et al.’s meta-analysis who estimate that a 15- ppb increase in indoor NO_2_ was associated with odds of current and lifetime asthma of 1.09 (95% CI 0.91–1.31). It is worth noting that this estimate draws on only four studies. In any case, using this point estimate would yield an increased risk of 6% of developing childhood asthma.

There are a number of limitations of this approach and assumptions necessary to make inferences. For example, we must assume here that the observed 48-hour differences in this study are steady over childhood, i.e., that the difference between a child growing up in a household that uses gas vs. the counterfactual scenario where that child grows up in a household using induction is always 9.88 ppb. We must also assume that the concentration-response function is linear. A limitation of this risk assessment approach is that the difference in mean 48-hour exposures we see between stove assignment are driven by extreme peaks in exposure (i.e., exposure contrasts between gas vs. induction during cooking exceed 9.88 ppb but average out given that when no cooking occurs differences are roughly 0). The implications of these spikes in short-term exposure differences for health risk are not self-evidence. In spite of these limitations, we think it useful to provide a benchmark for implied health risk from observed effect sizes in this study.

#### Short-term mortality

We parallel our above approach by drawing on Meng et al.’s estimate that, on average, a 10 *μgm*^−3^ increase in previous day ambient NO2 concentration was associated with a 0.46% (95% confidence interval 0.36%–0.57%) increase in all-cause mortality. So, we convert 10 *μgm*^−3^ to 5.32 ppb. Then, we multiply 0.46% by 9.88 divided by 5.32. Thus, we get an estimate of an increase of 0.85% in next day mortality.

Similar to above, some assumptions are required for this estimate. Namely, we assume that the concentration-response function is linear and that differences are constant (here, only for a day).

---

**Table S1.**
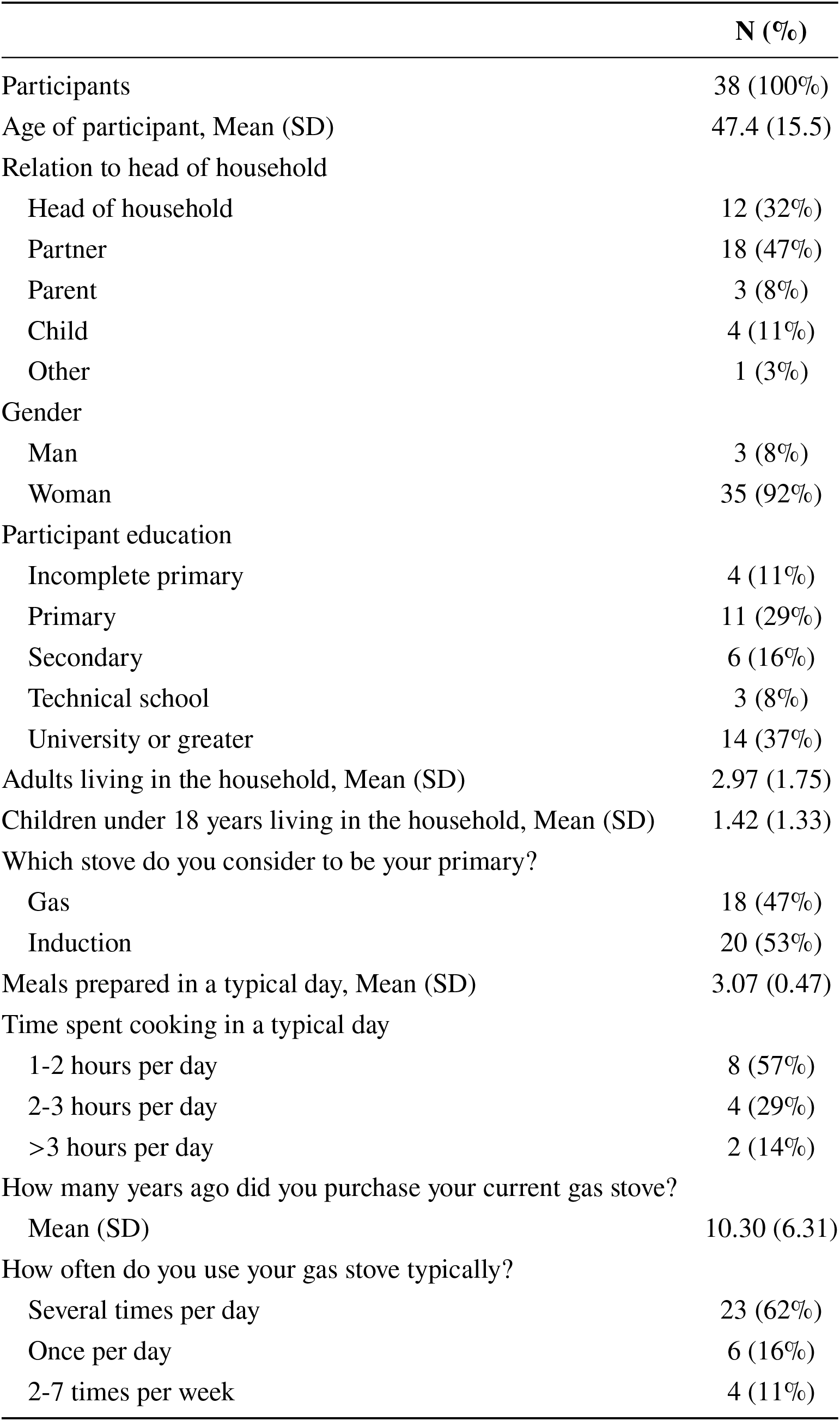
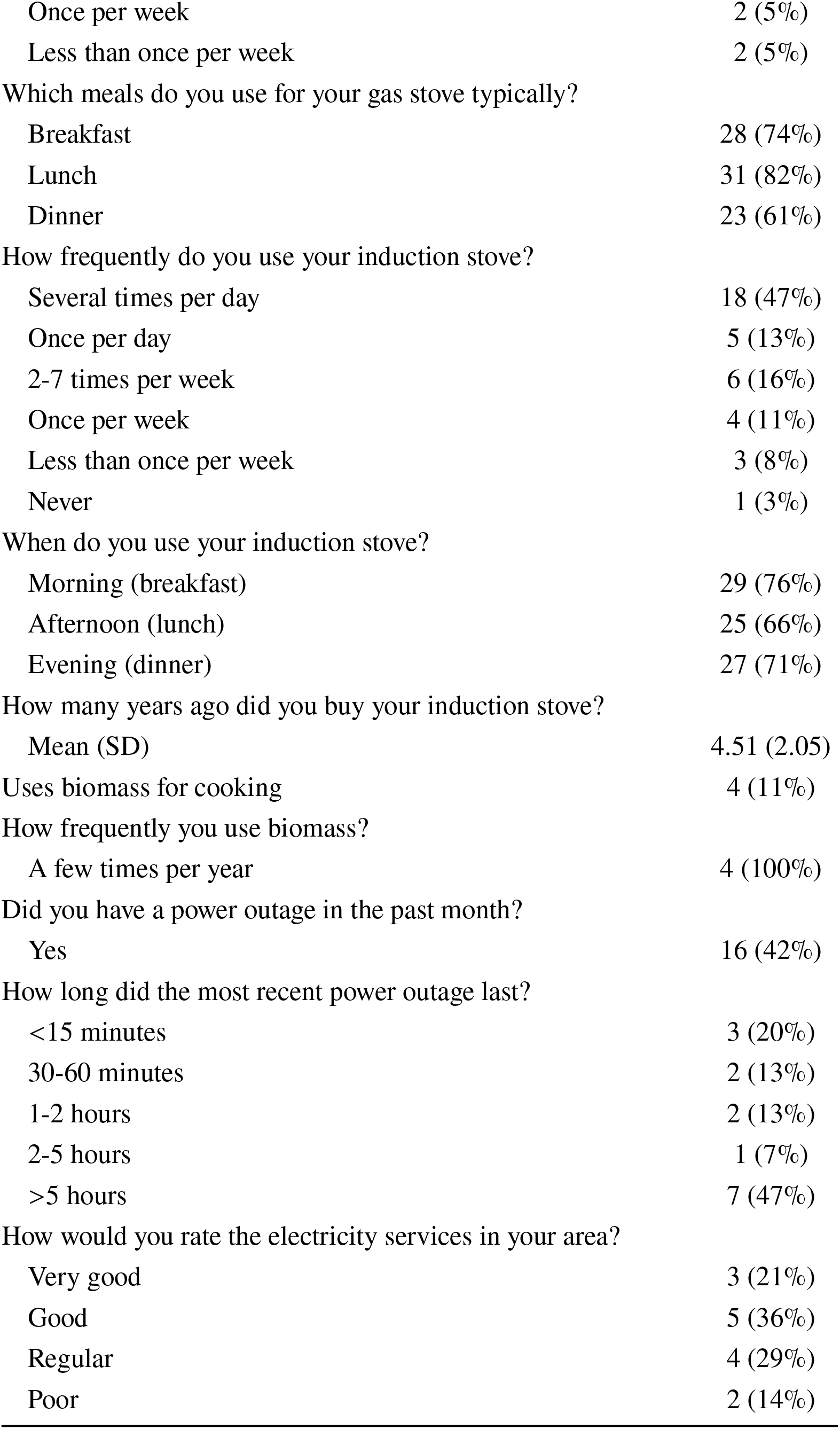
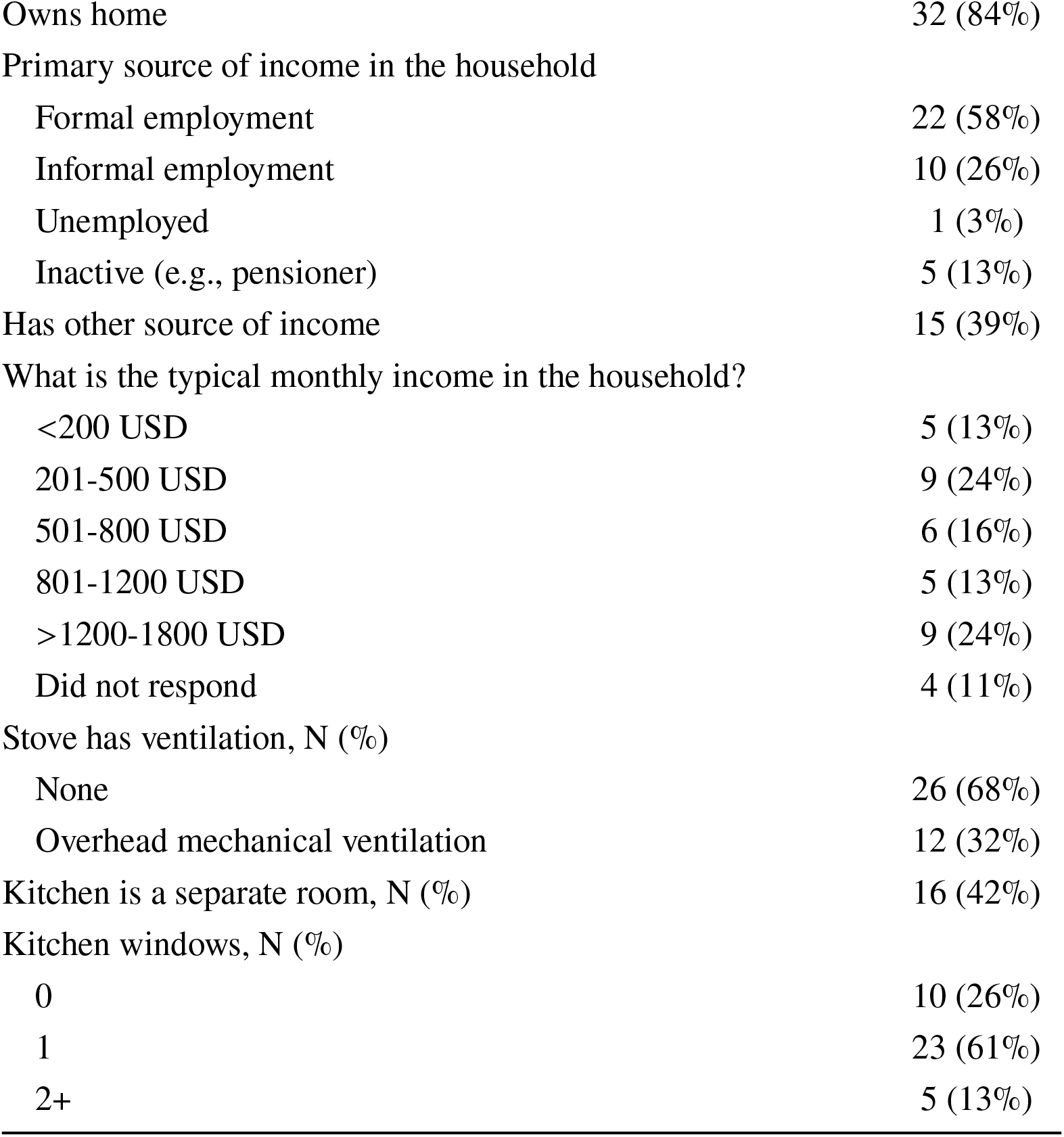
Participant and household characteristics.

|  | N (%) |
| --- | --- |
| Participants | 38 (100%) |
| Age of participant, Mean (SD) | 47.4 (15.5) |
| Relation to head of household |  |
| Head of household | 12 (32%) |
| Partner | 18 (47%) |
| Parent | 3 (8%) |
| Child | 4 (11%) |
| Other | 1 (3%) |
| Gender |  |
| Man | 3 (8%) |
| Woman | 35 (92%) |
| Participant education |  |
| Incomplete primary | 4 (11%) |
| Primary | 11 (29%) |
| Secondary | 6 (16%) |
| Technical school | 3 (8%) |
| University or greater | 14 (37%) |
| Adults living in the household, Mean (SD) | 2.97 (1.75) |
| Children under 18 years living in the household, Mean (SD) | 1.42 (1.33) |
| Which stove do you consider to be your primary? |  |
| Gas | 18 (47%) |
| Induction | 20 (53%) |
| Meals prepared in a typical day, Mean (SD) | 3.07 (0.47) |
| Time spent cooking in a typical day |  |
| 1-2 hours per day | 8 (57%) |
| 2-3 hours per day | 4 (29%) |
| >3 hours per day | 2 (14%) |
| How many years ago did you purchase your current gas stove? |  |
| Mean (SD) | 10.30 (6.31) |
| How often do you use your gas stove typically? |  |
| Several times per day | 23 (62%) |
| Once per day | 6 (16%) |
| 2-7 times per week | 4 (11%) |
| Once per week | 2 (5%) |
| Less than once per week | 2 (5%) |
| Which meals do you use for your gas stove typically? |  |
| Breakfast | 28 (74%) |
| Lunch | 31 (82%) |
| Dinner | 23 (61%) |
| How frequently do you use your induction stove? |  |
| Several times per day | 18 (47%) |
| Once per day | 5 (13%) |
| 2-7 times per week | 6 (16%) |
| Once per week | 4 (11%) |
| Less than once per week | 3 (8%) |
| Never | 1 (3%) |
| When do you use your induction stove? |  |
| Morning (breakfast) | 29 (76%) |
| Afternoon (lunch) | 25 (66%) |
| Evening (dinner) | 27 (71%) |
| How many years ago did you buy your induction stove? |  |
| Mean (SD) | 4.51 (2.05) |
| Uses biomass for cooking | 4 (11%) |
| How frequently you use biomass? |  |
| A few times per year | 4 (100%) |
| Did you have a power outage in the past month? |  |
| Yes | 16 (42%) |
| How long did the most recent power outage last? |  |
| <15 minutes | 3 (20%) |
| 30-60 minutes | 2 (13%) |
| 1-2 hours | 2 (13%) |
| 2-5 hours | 1 (7%) |
| >5 hours | 7 (47%) |
| How would you rate the electricity services in your area? |  |
| Very good | 3 (21%) |
| Good | 5 (36%) |
| Regular | 4 (29%) |
| Poor | 2 (14%) |
| Owns home | 32 (84%) |
| Primary source of income in the household |  |
| Formal employment | 22 (58%) |
| Informal employment | 10 (26%) |
| Unemployed | 1 (3%) |
| Inactive (e.g., pensioner) | 5 (13%) |
| Has other source of income | 15 (39%) |
| What is the typical monthly income in the household? |  |
| <200 USD | 5 (13%) |
| 201-500 USD | 9 (24%) |
| 501-800 USD | 6 (16%) |
| 801-1200 USD | 5 (13%) |
| >1200-1800 USD | 9 (24%) |
| Did not respond | 4 (11%) |
| Stove has ventilation, N (%) |  |
| None | 26 (68%) |
| Overhead mechanical ventilation | 12 (32%) |
| Kitchen is a separate room, N (%) | 16 (42%) |
| Kitchen windows, N (%) |  |
| 0 | 10 (26%) |
| 1 | 23 (61%) |
| 2+ | 5 (13%) |

**Table S2:** Descriptive statistics of study variables.

|  | Overall (N=76) | LPG (N=38) | Induction (N=38) |
| --- | --- | --- | --- |
| Randomly assigned as first period |  | 18 | 20 |
| Ambient 48-hour mean NO2 concentrations, ppb |  |  |  |
| Observations | 76 | 38 | 38 |
| Mean (SD) | 16.24 (5.43) | 16.89 (5.80) | 15.58 (5.02) |
| Median (IQR) | 16.13 (14.18, 19.11) | 17.29 (14.98, 20.51) | 15.32 (13.83, 18.37) |
| Ambient 48-hour mean PM2.5 concentrations, $\mu\text{gm}^{-3}$ | | | |
| Observations | 76 | 38 | 38 |
| Mean (SD) | 13.41 (2.58) | 13.42 (2.46) | 13.41 (2.73) |
| Median (IQR) | 13.25 (11.56, 14.95) | 12.94 (12.02, 14.84) | 13.57 (10.93, 15.14) |
| Mean 48-hour personal NO2 exposure, ppb |  |  |  |
| Observations | 76 | 38 | 38 |
| Mean (SD) | 19.76 (12.28) | 24.52 (14.86) | 15.00 (6.18) |
| Median (IQR) | 17.90 (12.37, 24.51) | 22.05 (17.57, 27.47) | 13.31 (10.73, 18.00) |
| Mean 48-hour kitchen area NO2 concentrations, ppb |  |  |  |
| Observations | 21 | 11 | 10 |
| Mean (SD) | 17.91 (3.00) | 18.63 (2.93) | 17.12 (3.03) |
| Median (IQR) | 18.84 (17.95, 19.86) | 19.75 (19.04, 20.05) | 18.33 (17.55, 18.52) |
| Mean 48-hour personal PM2.5 exposure, $\mu\text{gm}^{-3}$ | | | |
| Observations | 25 | 12 | 13 |
| Mean (SD) | 24.91 (16.45) | 30.21 (20.24) | 20.02 (10.55) |
| Median (IQR) | 22.46 (15.80, 31.18) | 23.76 (20.62, 33.54) | 17.76 (13.73, 27.75) |
| Personal PM2.5 monitor temperature, degrees C |  |  |  |
| Observations | 74 | 37 | 37 |
| Mean (SD) | 21.20 (1.87) | 21.24 (1.82) | 21.17 (1.94) |
| Median (IQR) | 21.45 (19.95, 22.62) | 21.42 (19.73, 22.70) | 21.64 (20.09, 22.47) |
| Detected personal PM2.5 monitor motion, minutes |  |  |  |
| Observations | 76 | 38 | 38 |
| Mean (SD) | 335.50 (188.78) | 352.33 (178.15) | 318.66 (199.80) |
| Median (IQR) | 343.50 (176.50, 441.75) | 377.00 (199.12, 436.75) | 294.50 (137.50, 446.75) |
| Detected LPG cooking, minutes |  |  |  |
| Observations | 38 | 38 | 38 |
| Mean (SD) | 279.42 (136.41) | 279.42 (136.41) | 7.95 (25.85) |
| Median (IQR) | 262.00 (172.75, 381.50) | 262.00 (172.75, 381.50) | 0.00 (0.00, 0.00) |
| Detected induction cooking, minutes |  |  |  |
| Observations | 38 | 38 | 38 |
| Mean (SD) | 284.37 (147.21) | 1.34 (8.27) | 284.37 (147.21) |
| Median (IQR) | 289.00 (161.25, 368.75) | 0.00 (0.00, 0.00) | 289.00 (161.25, 368.75) |

**Table S3:**
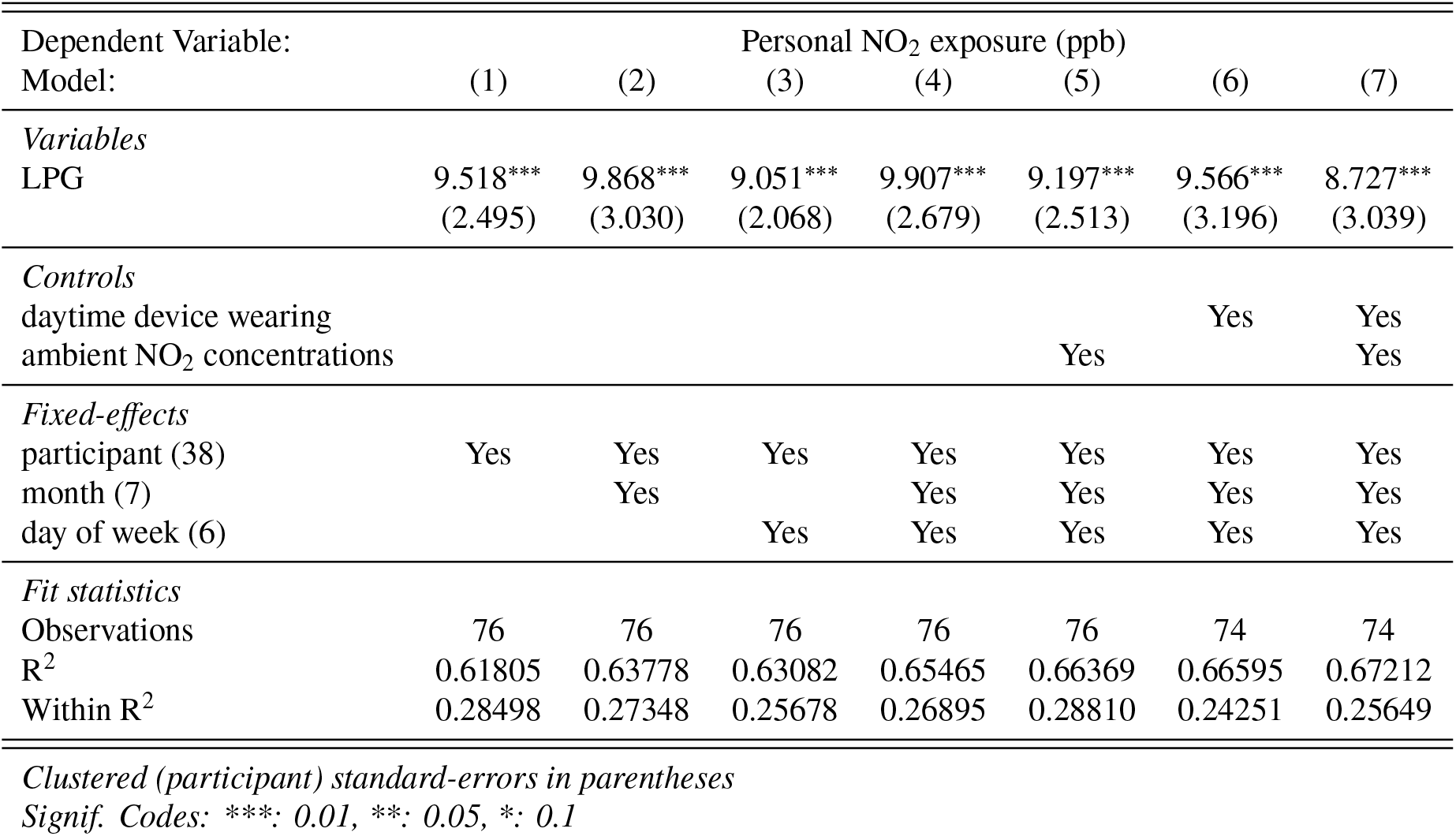
Association between assignment to using the LPG stove and two-day average personal NO_2_ exposure.

**Table S4:**
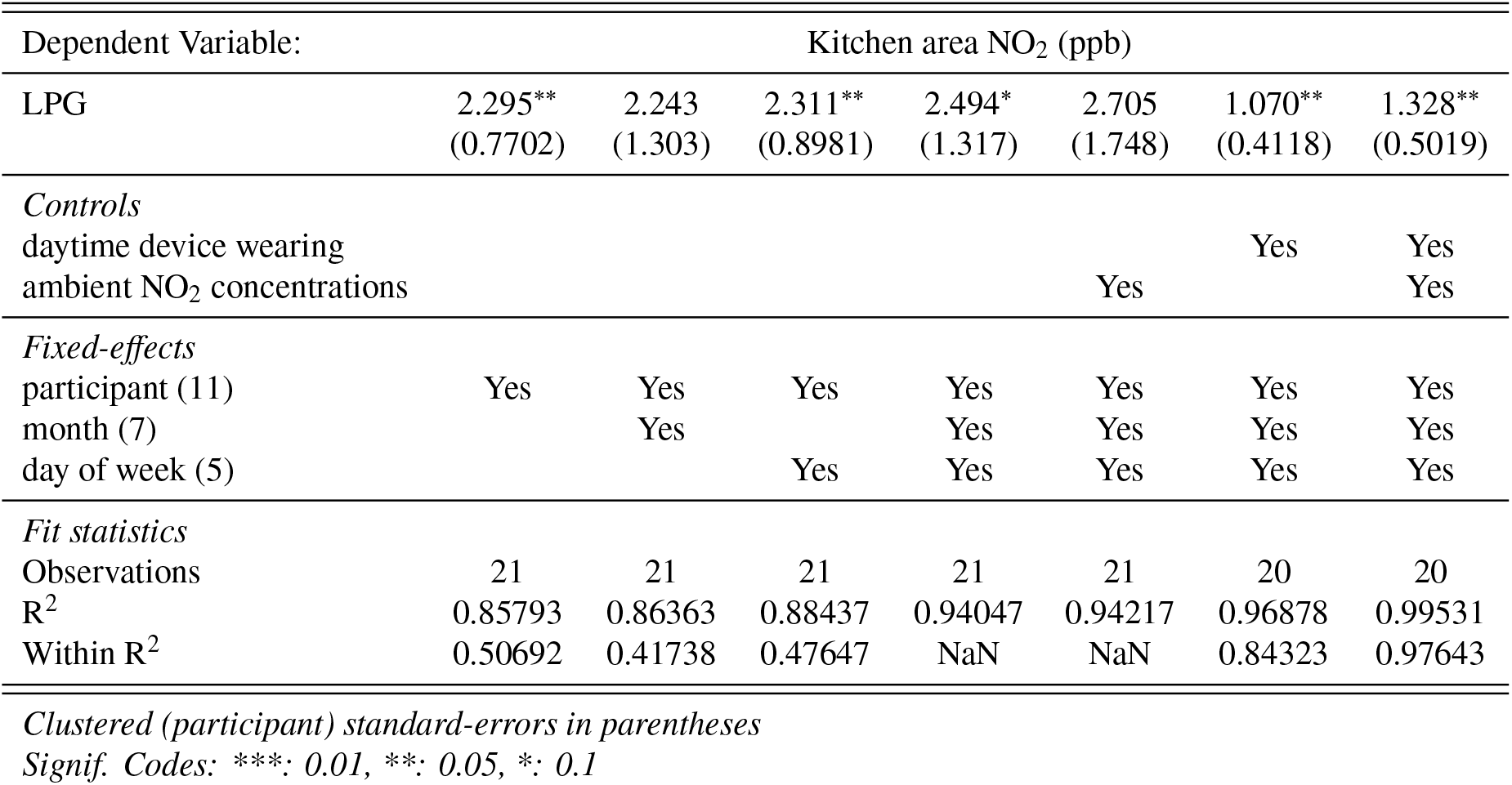
Association between assignment to using the LPG stove and two-day average kitchen area NO_2_ concentrations.

| Dependent Variable: | Kitchen area NO<sub>2</sub> (ppb) |  |  |  |  |  |  |
| --- | --- | --- | --- | --- | --- | --- | --- |
| LPG | 2.295**<br>(0.7702) | 2.243<br>(1.303) | 2.311**<br>(0.8981) | 2.494*<br>(1.317) | 2.705<br>(1.748) | 1.070**<br>(0.4118) | 1.328**<br>(0.5019) |
| *Controls* |  |  |  |  |  |  |  |
| daytime device wearing |  |  |  |  |  | Yes | Yes |
| ambient NO<sub>2</sub> concentrations |  |  |  |  | Yes |  | Yes |
| *Fixed-effects* |  |  |  |  |  |  |  |
| participant (11) | Yes | Yes | Yes | Yes | Yes | Yes | Yes |
| month (7) |  | Yes |  | Yes | Yes | Yes | Yes |
| day of week (5) |  |  | Yes | Yes | Yes | Yes | Yes |
| *Fit statistics* |  |  |  |  |  |  |  |
| Observations | 21 | 21 | 21 | 21 | 21 | 20 | 20 |
| R<sup>2</sup> | 0.85793 | 0.86363 | 0.88437 | 0.94047 | 0.94217 | 0.96878 | 0.99531 |
| Within R<sup>2</sup> | 0.50692 | 0.41738 | 0.47647 | NaN | NaN | 0.84323 | 0.97643 |

*Clustered (participant) standard-errors in parentheses*
*Signif. Codes: \*\*\*: 0.01, \*\*: 0.05, \*: 0.1*

**Table S5:** Association between assignment to using the LPG stove and two-day average personal PM_2.5_ exposure.

| Dependent Variable:<br>Model: | (1) | (2) | (3) | Personal PM<sub>2.5</sub> exposure ($\mu\text{gm}^{-3}$) | | | | | |
| --- | --- | --- | --- | --- | --- | --- | --- | --- | --- |
|  |  |  |  | (4) | (5) | (6) | (7) | (8) | (9) |
| *Variables* |  |  |  |  |  |  |  |  |  |
| LPG | 11.46*<br>(5.997) | 12.71<br>(8.171) | 8.719*<br>(4.600) | 11.35*<br>(5.340) | 12.52*<br>(6.439) | 13.88<br>(8.604) | 9.974<br>(7.062) | 13.72*<br>(7.252) | 13.24*<br>(6.950) |
| *Controls* |  |  |  |  |  |  |  |  |  |
| daytime device wearing |  |  |  |  |  | Yes | Yes |  |  |
| ambient NO<sub>2</sub> concentrations |  |  |  |  | Yes |  | Yes |  |  |
| *Fixed-effects* |  |  |  |  |  |  |  |  |  |
| participant (13) | Yes | Yes | Yes | Yes | Yes | Yes | Yes | Yes | Yes |
| month (7) |  | Yes |  | Yes |  | Yes |  |  |  |
| day of week (5) |  |  | Yes | Yes |  |  | Yes |  |  |
| *Fit statistics* |  |  |  |  |  |  |  |  |  |
| Observations | 25 | 25 | 25 | 25 | 25 | 25 | 25 | 24 | 24 |
| R<sup>2</sup> | 0.64716 | 0.66373 | 0.73694 | 0.79814 | 0.69604 | 0.71843 | 0.78134 | 0.65662 | 0.69466 |
| Within R<sup>2</sup> | 0.25589 | 0.27013 | 0.09539 | 0.16426 | 0.35897 | 0.38885 | 0.24806 | 0.28942 | 0.36814 |
| *Clustered (participant) standard-errors in parentheses* |  |  |  |  |  |  |  |  |  |
| *Signif. Codes: \*\*\*: 0.01, \*\*: 0.05, \*: 0.1* |  |  |  |  |  |  |  |  |  |

**Table S6:**
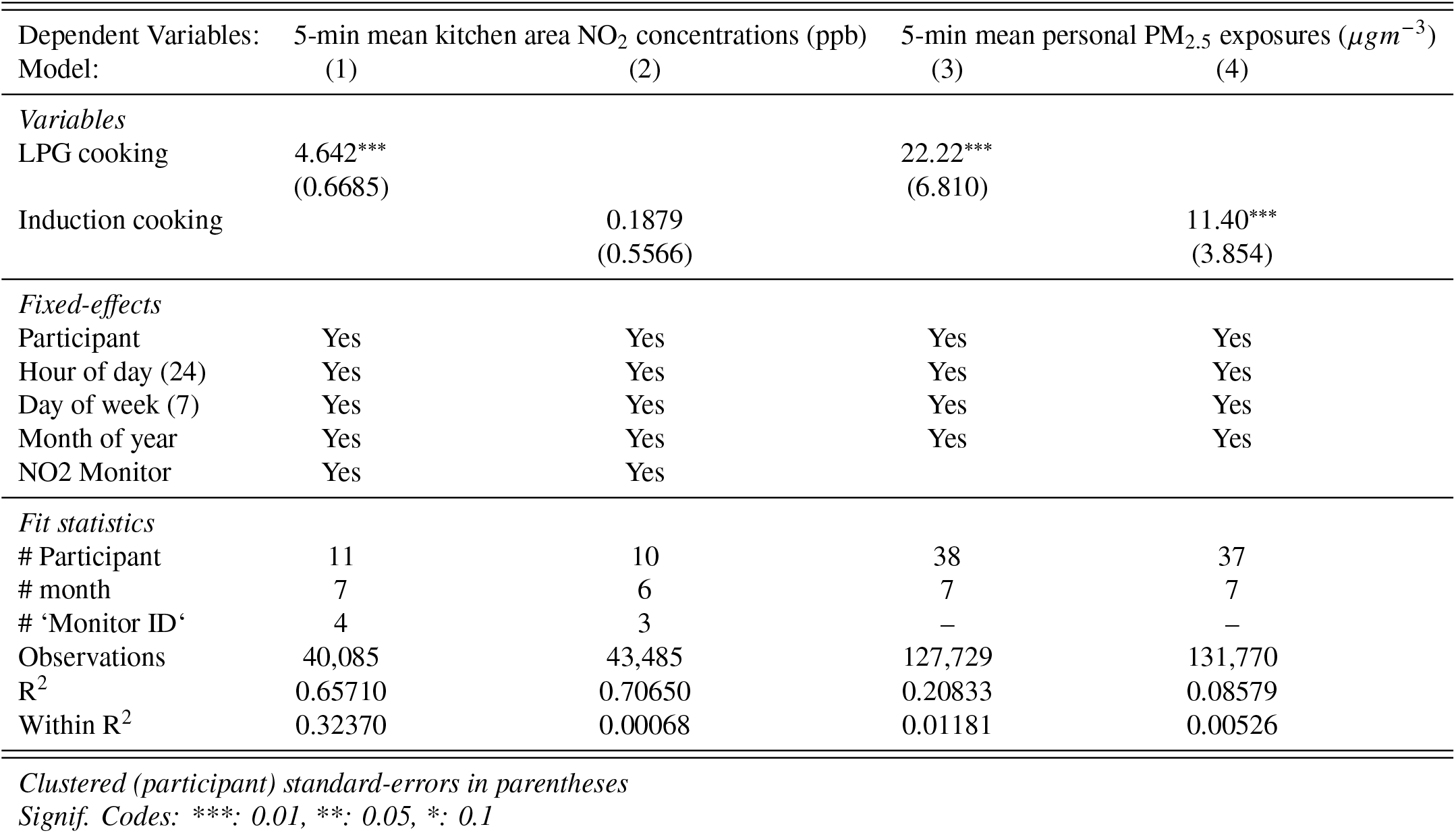
Association between detected cooking events and 5-minute average air pollution concentrations.

**Table S7:** Effect of cooking on pollution during sessions with both induction and LPG stove monitoring.

| Dependent Variables: | 5-min mean kitchen area NO<sub>2</sub> concentrations (ppb) | 5-min mean personal PM<sub>2.5</sub> exposures ($\mu\text{gm}^{-3}$) |
| --- | --- | --- |
| Model: | (1) | (2) |
| *Variables* |  |  |
| Induction cooking | -0.3957**<br>(0.1424) | 3.814<br>(2.303) |
| LPG cooking | 4.193***<br>(0.5053) | 30.87**<br>(13.06) |
| *Fixed-effects* |  |  |
| Participant | Yes | Yes |
| Hour of day (24) | Yes | Yes |
| Day of week (7) | Yes | Yes |
| Month of year (6) | Yes | Yes |
| NO2 Monitor (2) | Yes |  |
| *Fit statistics* |  |  |
| # Participants | 7 | 15 |
| Observations | 55,944 | 46,369 |
| R<sup>2</sup> | 0.50035 | 0.13604 |
| Within R<sup>2</sup> | 0.34363 | 0.02924 |

*Clustered (participant) standard-errors in parentheses*
*Signif. Codes: \*\*\*: 0.01, \*\*: 0.05, \*: 0.1*

**Figure S2:**
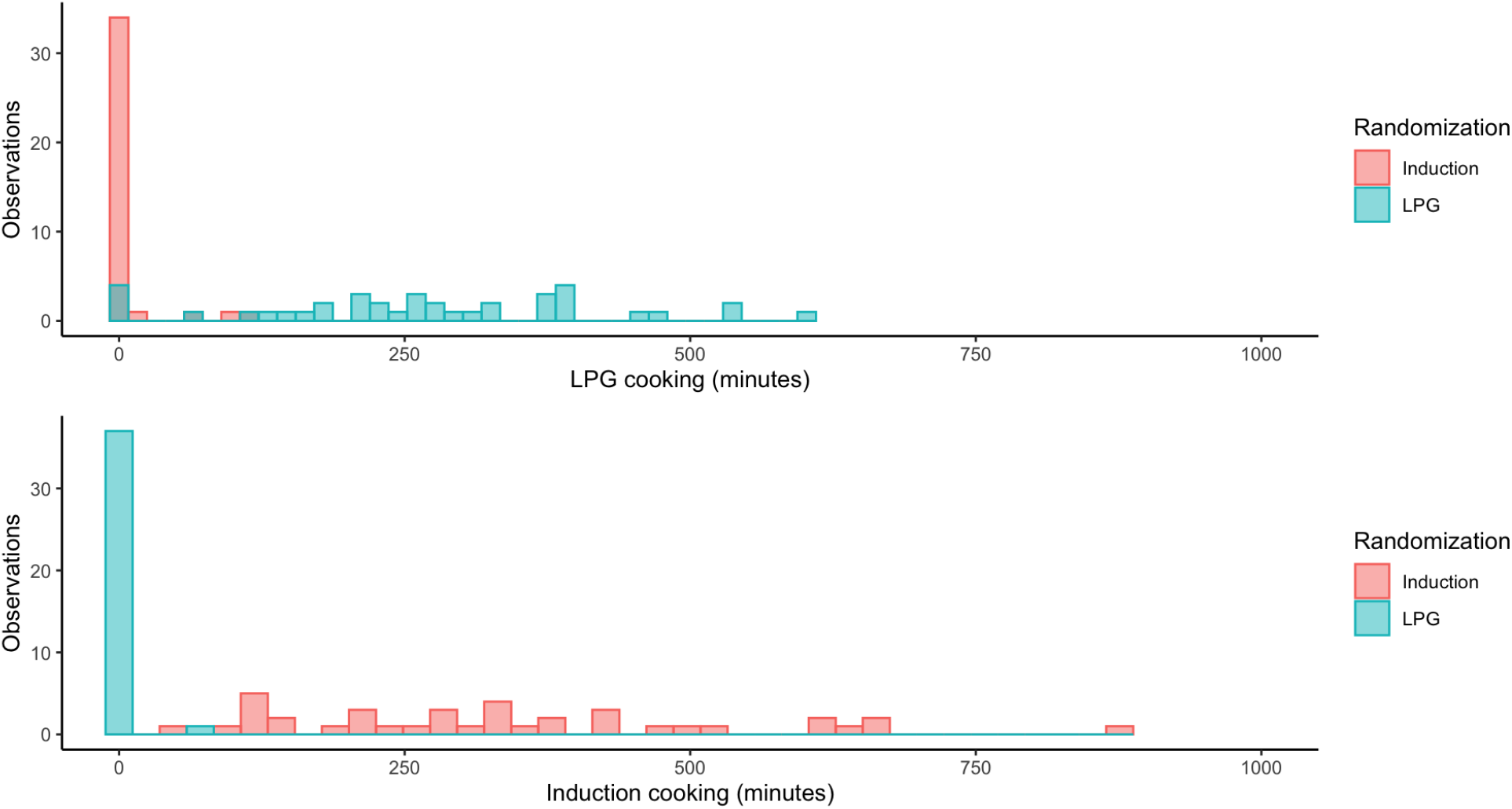
Minutes of detected LPG and induction stove use by randomization. In total, we observed deviations from assigned stove use during 4.4% of total cooking time when LPG and induction stove use were measured (272 out of 6243 minutes across 15 monitoring sessions).

**Figure S3:**
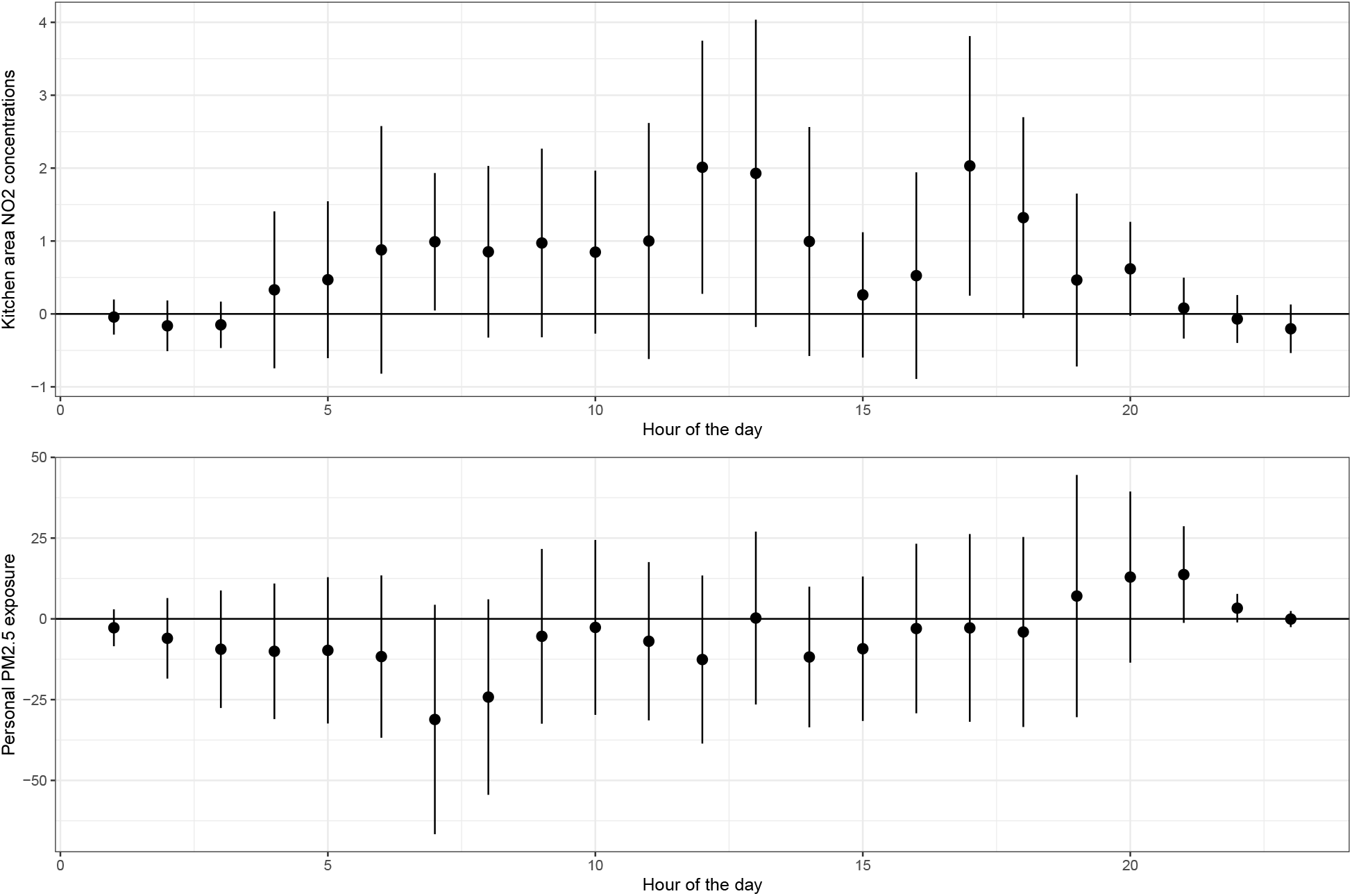
Hour-wise estimates of the difference between kitchen area NO2 concentrations and personal PM2.5 exposures when households were randomized to use gas vs. induction. Coefficients and 95% confidence intervals are displayed from a regression that interacts randomized stove assigned with hour of the day, controlling for participant, day of week, and month fixed effects (the NO2 regression accounts for monitor FE). Standard errors clustered at participant level. Estimates that are above zero indicate that average pollution was higher when the household was randomized to use gas.

## Notes

**Conflicts of Interest** The authors declare they have nothing to disclose.

### Competing Interest Statement

The authors have declared no competing interest.

### Author Declarations

Research was approved by the Institutional Review Boards at the Columbia University Medical Center and the Bioethics Committee at the Universidad San Francisco de Quito (USFQ). USFQ approved COVID-19 safety protocols for in-person activities.

